# On the robustness of medical term representations in locally deployable language models

**DOI:** 10.64898/2026.02.24.26346972

**Authors:** Stephen D. Auger, Neil S.N. Graham, Gregory Scott

## Abstract

**Background:** Hosting large language models (LLMs) on-premises can secure patient data but requires compact architectures to function on standard hardware. The impact of such constraints on the robustness of their representations for medical terminology is important for clinical AI safety but poorly understood. The statistical nature of LLM training inherently limits the representation of terms with low societal prominence or lexical frequency, and high ambiguity.

**Methods:** We assessed 15 open-weights LLMs (4B–120B) for their representational robustness of 250 neurological terms. Neurology was chosen for its strict hierarchical and anatomical terminology. A term’s representation was deemed robust only if the model correctly navigated four tests, verifying valid links against distractors and reverse associations. We examined associations between representational robustness and model size, medical fine-tuning, and five terminological subdomains (localisation, clinical features, investigations, diagnoses, and treatments). We assessed term difficulty using the semantic complexity index (SCI), a novel composite integrating societal prominence, lexical frequency, and ambiguity.

**Results:** Representational robustness followed a log-linear scaling law relative to model size (r=0.736, p=0.002). Medical fine-tuning yielded no benefit for 4B models, but significantly improved larger 27B model performance, with rate of robust representations rising from 38.2% to 62.6% (p<0.0001). While most local-LLMs’ performance degraded sharply with increasing SCI values, GPT-OSS 20B and 120B maintained ‘complexity invariance’ (with <20% decline from lowest to highest complexity terms). Notably, the general-purpose 20B GPT-OSS model outperformed larger and medically fine-tuned counterparts. Robustness varied by subdomain (F=4.69, p=0.003), with diagnoses (73.8%) scoring significantly higher than localisation (47.9%, p=0.004) and clinical features (52.1%, p=0.02).

**Conclusions:** While representational robustness broadly follows model size scaling laws, neither model size nor fine-tuning guarantees clinical reliability. Since performance fluctuates with terminological complexity and subdomain, safe deployment requires validating representational robustness for specific use cases rather than assuming larger models handle medical language safely.

**Sentence Description:** This study shows that model size and medical fine-tuning are not reliable indicators of clinical robustness across 15 locally-deployable LLMs. Because performance varies significantly by terminological complexity and subdomain, safe application requires validation methods that account for these factors.

## INTRODUCTION

Hosting large language models (LLMs) on-premises promises a compelling solution for healthcare and research organisations looking to balance AI adoption with data privacy requirements^1^. Local deployment allows organisations to adhere to strict governance frameworks like HIPAA^2^ and GDPR^3^ while ensuring operational resilience during network outages^4,5^. This local approach generally necessitates the use of lightweight LLMs able to run on the hardware available in these settings^6–9^. However, it is unclear if there is a threshold at which the operational requirement for smaller LLMs compromises their safety for clinical use.

While lightweight LLMs often retain convincing linguistic fluency, this surface-level capability may not guarantee the robust representation of medical terminology that is likely to be a prerequisite for safe clinical deployment^10^. That is, a model may generate statistically probable text linking two medical terms while having only a partial representation of the underlying relationship between them^11,12^. While one might assume that larger LLMs would contain more robust representations of medical terminology, for example, that a 70 billion parameter (70B) model would outperform 20B models, this is unproven.

Another approach to improving the robustness of LLM representations is via fine-tuning, i.e., additional training with specialist data^13^. However, the relative gains in representational robustness from increasing model size versus medical fine-tuning are unclear.

Here, we evaluated the robustness of a range of locally deployable LLMs’ representations of medical terminology and explored their suitability for different clinical use cases. Clinical neurology was selected as the testing domain due to its reliance on complex nomenclature and rigid conceptual hierarchies^14^, both of which necessitate robust terminological representations.

To test the robustness of LLMs’ representations, rather than use simple multiple-choice questions (e.g. MedQA-USMLE and other similar benchmarks)^6,15–17^, we employed a novel method that probed models’ representations of the relationships between terms. We then tested how models’ representational robustness varied with their parameter size and the use of medical fine-tuning. We additionally explored how representational robustness varied according to a composite measure of the societal prominence, ambiguity, and lexical properties of terms. We also examined the relative performance of models across five subdomains of terms relating to anatomical localisation, clinical features (symptoms or signs), investigations, diagnoses, and treatments. In doing so, we aim to provide evidence to inform healthcare organisations and researchers looking to deploy lightweight clinical LLMs, as well as a framework for assessing model suitability for specific use cases.

## METHODS

### Test dataset creation and definition of a ‘robust representation’ of medical terms

We created a dataset of 250 clinical neurology *term triplets* to use for evaluating the robustness of terminological representations within a range of lightweight LLMs. Each triplet took the form: *child term [A], parent category [B], distractor [C]*. For a specific term to be classified as robustly represented, an LLM was required to correctly identify four logical relationships involving the term’s triplet A, B, and C. (Figure 1A). For example, given the term triplet (Miller-Fisher syndrome, Guillain-Barré variant, Charcot-Marie-Tooth variant), the LLM was asked to: (1) Affirm that B is a parent of A, i.e., “Miller-Fisher syndrome guarantees presence of a variant of Guillain-Barré syndrome” is true; (2) Reject that A is the parent of B, i.e., “Variant of Guillain-Barré syndrome guarantees presence of Miller-Fisher syndrome” is false (3) Distinguish the term from a clinically distinct distractor, i.e., “Miller-Fisher syndrome guarantees presence of a variant of Charcot-Marie-Tooth disease” is false, and (4) Reject the reverse implication from the distractor i.e., “Variant of Charcot-Marie-Tooth disease guarantees presence of Miller-Fisher syndrome” is false.

**Figure 1.**
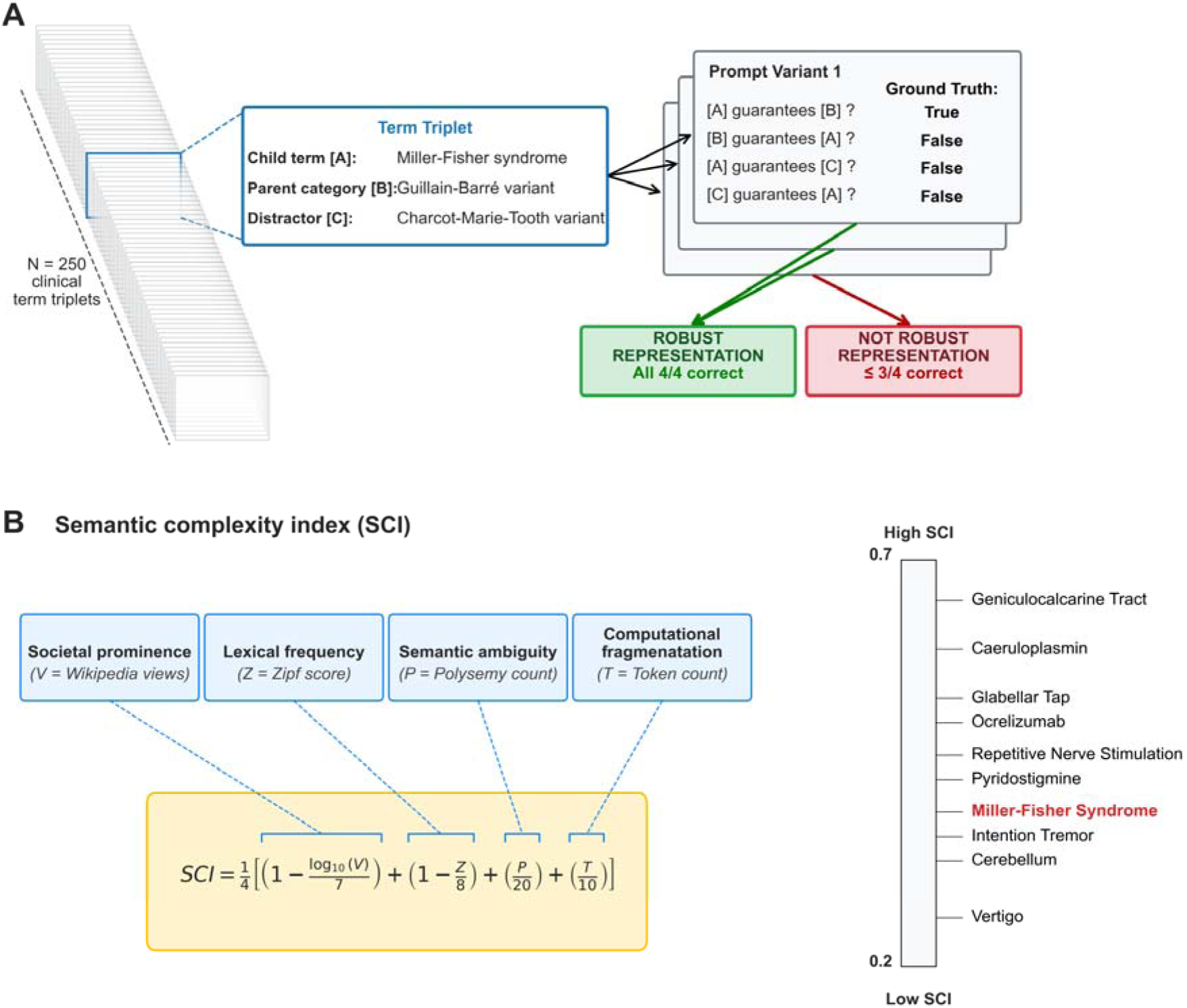
(A) Assessment of LLM representational robustness. Models were evaluated with 250 clinical terms (child terms [A]) based on their ability to correctly identify four logical relationships involving a parent category term [B] and a distractor term [C]. A term was classified as “robustly represented” only if the model correctly answered all four questions within a term triplet. Each triplet was tested across three prompt variants, totalling 750 unique evaluations per model. **(B) Semantic complexity index (SCI).** To analyse how linguistic properties influence model performance, we developed the SCI, a composite metric grading the relative difficulty of medical terms (range, 0.2 [lowest complexity] to 0.7 [highest complexity]). The index integrates four variables: societal prominence (annual English Wikipedia views), lexical frequency (Zipf score), semantic ambiguity (polysemy), and computational fragmentation (token count).

An LLM was defined as having a “robust representation” of a term if it correctly answered the four logical relationships involving the term’s triplet. Passing this standard differentiates models that encode the precise directional nature of relationships from those relying on general statistical association. Note that a model relying on random chance has only a 6.25% (1/16) probability of passing a term triplet (i.e., making four consecutive binary selections each with probability ½).

The ground truth of the four logical relationships for each term triplet was validated by three Neurologists on the UK Specialist Register (SDA, NSNG, GS).

To control for prompt fragility, we included three prompt variations to query the directional relationships (see Supplementary Appendix). We employed a strict zero-shot protocol, explicitly excluding few-shot examples to prevent any in-context learning bias. This yielded 750 unique term evaluations (250 triplets × 3 prompt variants) per model. We defined the ‘robust representation rate’ as the percentage of these 750 term evaluations answered correctly.

### Semantic complexity index

To evaluate how the intrinsic properties of medical terms influence the robustness of their LLM representations, we developed a composite semantic complexity index (SCI) (Figure 1B). The SCI incorporates four data-derived components, each normalised to a 0–1 scale: (1) Societal prominence, measured using annual English Wikipedia views (V), log_10_-transformed to account for power-law distributions in web traffic data ^18,19^, and normalised against a fixed theoretical maximum of 7.0 (corresponding to the order of magnitude of the highest-traffic public pages’ 10^7^ annual views). (2) Lexical rarity, derived from Zipf frequency scores (Z)^20,21^, normalised against a maximum of 8 (the practical upper limit for highest-frequency words in natural language). (3) Semantic ambiguity, quantified as the number of different meanings (senses) for a term in WordNet (P)^22^. We capped and normalised this count to 20 to prevent rare, multi-meaning English verbs from ‘compressing’ the data; this ensures the index remains sensitive to the smaller, more relevant differences in ambiguity found between medical terms. (4) Computational fragmentation, defined by token count (T), relative to a limit of 10 tokens. The index was calculated as the mean of these components, with prominence and rarity inverted so that lower frequency increases the complexity score:

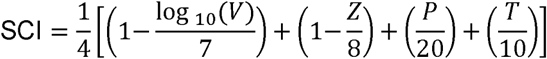

We normalised all components against fixed theoretical maximums so the SCI can serve as a transferable standard; low values imply lower representational challenge, while higher values imply specialised or ambiguous concepts that pose a more significant challenge for consistent LLM modelling.

### Five clinical subdomains

To evaluate whether representational robustness varies by clinical context, we constructed the test dataset to be balanced dataset across five distinct subdomains (n=50 each): anatomical localisation, clinical features, diagnoses, investigations, and treatments. We controlled for potential confounding by ensuring baseline semantic complexity (SCI) did not differ significantly between subdomains; this ensures that observed differences in robustness do not reflect variations in term-level difficulty.

### Model selection

We selected for evaluation 15 ‘lightweight’ open-weights LLMs capable of local ‘on-premises’ deployment. The details of the 15 LLMs are included in Table S1. These LLMs ranged from 4 billion to 120 billion parameters in size and represented distinct architectural families from providers including Google^23^, OpenAI^24^, Meta^25^, Mistral^26^, Alibaba^27^, Baidu^28^, Microsoft^29^, Cohere^30^, and DeepSeek^31^. As well as general-purpose LLMs, we considered variants with medical fine-tuning. We therefore included two MedGemma^32^ models in the final set of 15 LLMs. Other models with medical fine-tuning, namely BioMistral 7B^33^ and Meditron (7B/70B)^34^ were not included due to persistent instruction-following failures. These models consistently produced unstructured narrative or repetitions instead of the required outputs. This inability to follow zero-shot constraints rendered their latent knowledge inaccessible.

In addition to testing these 15 LLM, we also confirmed that the terms were linguistically representable by a transformer architecture, and accurate responses were possible with the specific prompts used, by testing responses from a frontier reference model, Google Gemini 3 Pro.

### Model execution

We evaluated each of the 15 LLMs, executing the 750 evaluations described above and calculating the robust representation rate. To ensure maximum stability and reproducibility of outputs, we set the temperature parameter to 0.0 for all models, while maintaining all other hyperparameters at their default configurations. We enforced realistic local deployment constraints by executing models on single-GPU instances (20GB–80GB VRAM), dynamically matching hardware specifications to the minimum operational requirements of each architecture.

### Statistics

95% confidence intervals (Cis) were calculated using the normal approximation to the binomial distribution, with variance derived from the dispersion of binary outcomes across 750 total observations (250 unique medical terms each evaluated across three distinct prompts).

We evaluated the performance of the 15 LLMs to assess how the robust representation rate varied according to four determinants.

First, we considered the impact of model parameter size by calculating the Pearson correlation between the robust representation rate and log-transformed LLM parameter counts^35^.

Second, we considered performance according to clinical subdomain using a one-way ANOVA (following verification of normality and homogeneity assumptions) followed by Tukey’s HSD post-hoc tests (α = 0.05) for significant main effects. To restrict analysis to models with meaningful discriminative signal, we excluded models performing at floor or ceiling levels (defined as SCI LOWESS-estimated accuracy of chance-level 6.25%, or ceiling 100% for the highest-complexity stimuli).

Third, to characterise the association between representation robustness and semantic complexity, we modelled the robust representation rate as a function of SCI using Locally Weighted Scatterplot Smoothing (LOWESS; with a standard smoothing fraction 0.66).

Fourth, we considered the effect of specialist medical fine-tuning. We compared standard Gemma 3 models against their medically fine-tuned counterparts, MedGemma (4B and 27B). To determine if fine-tuning was equally effective across different levels of term complexity, we stratified terms into two complexity cohorts using a median split of SCI scores. We evaluated the absolute efficacy of fine-tuning within each complexity cohort using Pearson’s Chi-squared test. Second, to test if fine-tuning yielded disproportionate gains for difficult terms, we analysed the interaction between model type and complexity. Specifically, for the 27B models, we performed a Z-test on the difference of differences to determine if the performance improvement for high-complexity terms was statistically distinct from that of low-complexity terms. All P-values were two-sided.

All statistical analyses were performed using Python (v3.13.5), using Pandas and NumPy for data management, SciPy and Statsmodels for statistical analysis, and Seaborn and Matplotlib for data visualisation.

## RESULTS

We evaluated 15 locally-deployable LLMs (4B-120B parameters) using the ‘robust representation rate’ for 750 neurological terms triplets (250 triplets x 3 prompt variants). A ‘pass’ was awarded only if a model correctly identified four directional relationships within a term triplet, including both positive connections and negative distractors (n=750).

The frontier reference model, Google Gemini Pro 3, established a 98.4% (±0.9% CI) robust representation rate ceiling (Figure 2), with rare errors resolving under alternative prompt phrasing. This confirms the feasibility of the evaluation task for LLMs and implies any performance differences with lightweight LLMs do not stem from inaccuracies or ambiguity in the prompts or data.

**Figure 2.**
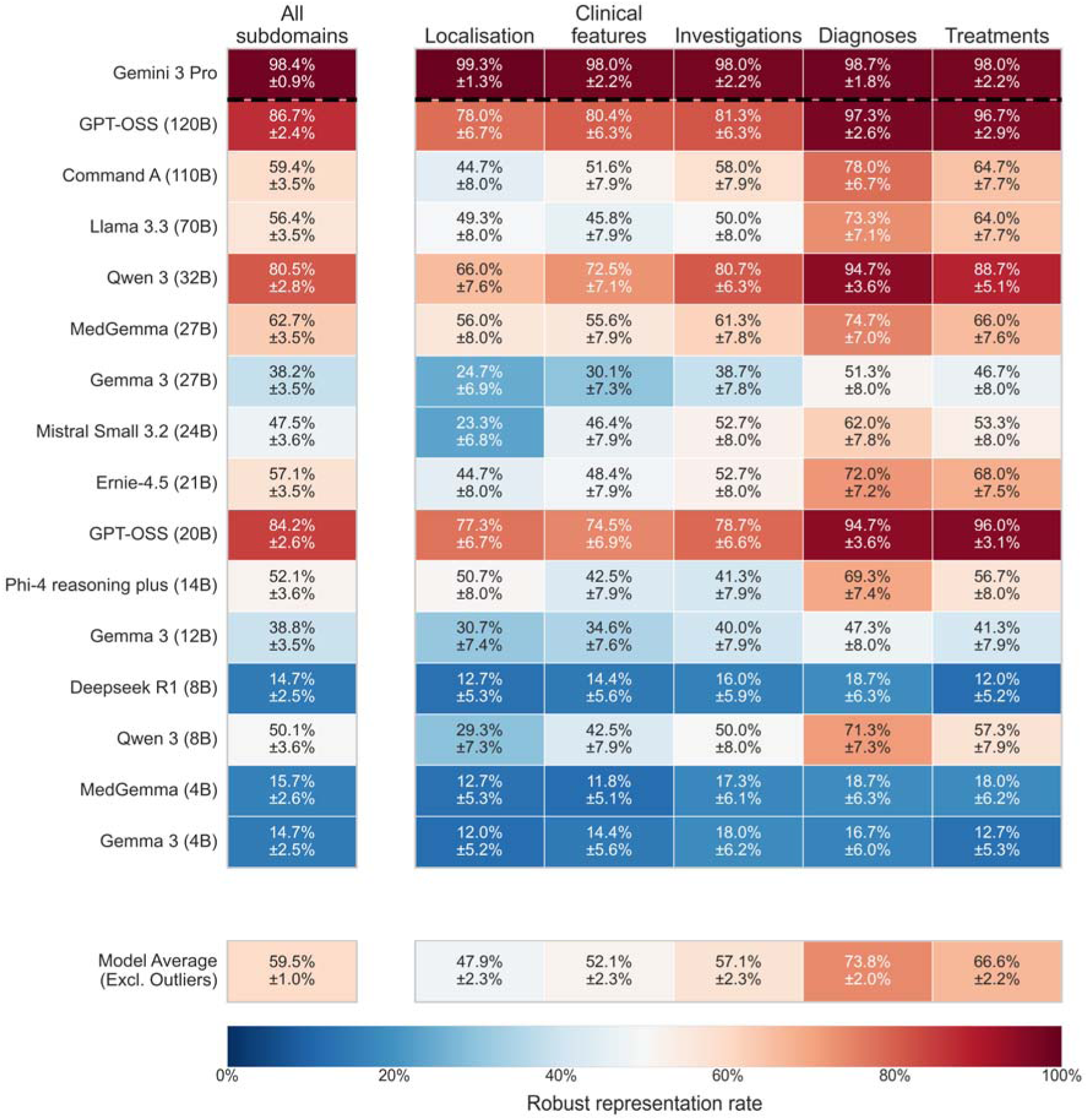
Representational robustness of medical terms across different models and clinical subdomains. This heatmap illustrates the robust representation rate (+/- 95% confidence interval). The data is visualised on a blue-red scale, where blue indicates low performance and red indicates high performance. Rows represent individual LLMs ordered from largest to smallest parameter size. Columns display performance broken down by five specific medical term categories. The left-most column shows a composite score, aggregating performance across all five subdomains for each model. The bottom row displays the average performance across all valid models for each of the five subdomains; models were excluded from this average value if their performance was at floor or ceiling level based on the LOWESS trajectory analysis of performance versus term complexity – specifically, models were omitted if their mean trajectory was at or below chance level (6.25%) or if they achieved 100% accuracy on highest complexity terms. Gemma 3 (4B), Deepseek R1 (8B) and Gemini Pro 3 were excluded by these criteria.

### Representational robustness scales with model size with notable exceptions

Performance ranged from 14.7% (± 2.5%) in the smallest models to 86.7% (± 2.4%) in the highest performing lightweight LLMs. While representational robustness was strongly correlated with model size (r = 0.736, p = 0.002), the data revealed significant departures from established log-linear LLM scaling laws^35^ (Figure 3). GPT-OSS (20B, 84.2% ± 2.6%) and Qwen 3 (32B, 80.5% ± 2.8%) demonstrated robustness rates comparable to or exceeding models of a substantially larger size (70B+).

**Figure 3.**
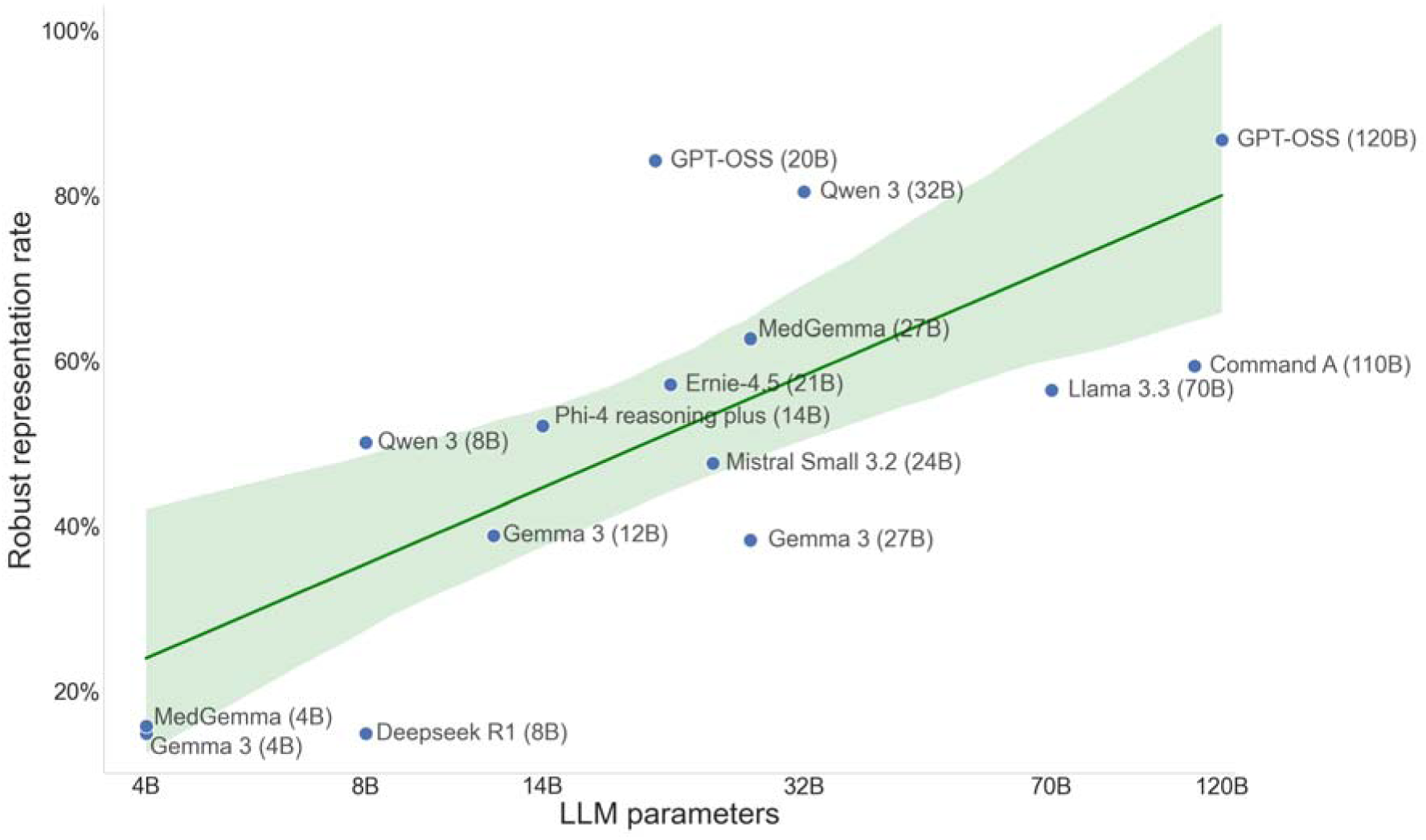
Model parameter size and representational robustness. The robust representation rate is plotted against model size on a logarithmic scale. The green line indicates a log-linear regression fit (shaded region = 95% confidence interval). While the data exhibits a general log-linear scaling law, where increased model size correlates with improved accuracy, clear deviations from the regression line highlight that additional factors influence performance at similar parameter counts.

### Representational robustness varies across clinical subdomains independent of semantic complexity

Representational robustness varied significantly across the five clinical subdomains (F=4.69, P=0.003), despite them being balanced according to SCI (F=2.24, P=0.07). Models demonstrated highest robustness for diagnostic terms (Figure 2, mean 73.8% ± 2.0%) and this was significantly superior to that for localisation (47.9% +/- 2.3%; mean difference 25.9%, adjusted P = 0.004) and clinical features (52.1% +/- 2.3%; mean difference 21.8%, adjusted P = 0.02). No other pairwise subdomain comparisons reached statistical significance after correction for family-wise error.

### Semantic complexity reveals divergent performance degradation profiles across models

To quantify the influence of linguistic complexity on representational robustness, we modelled the robust representation rate as a function of the SCI (incorporating the rarity, ambiguity, and fragmentation of terms). We identified distinct profiles of performance degradation with increasing semantic complexity across the evaluated models (Figure 4).

**Figure 4.**
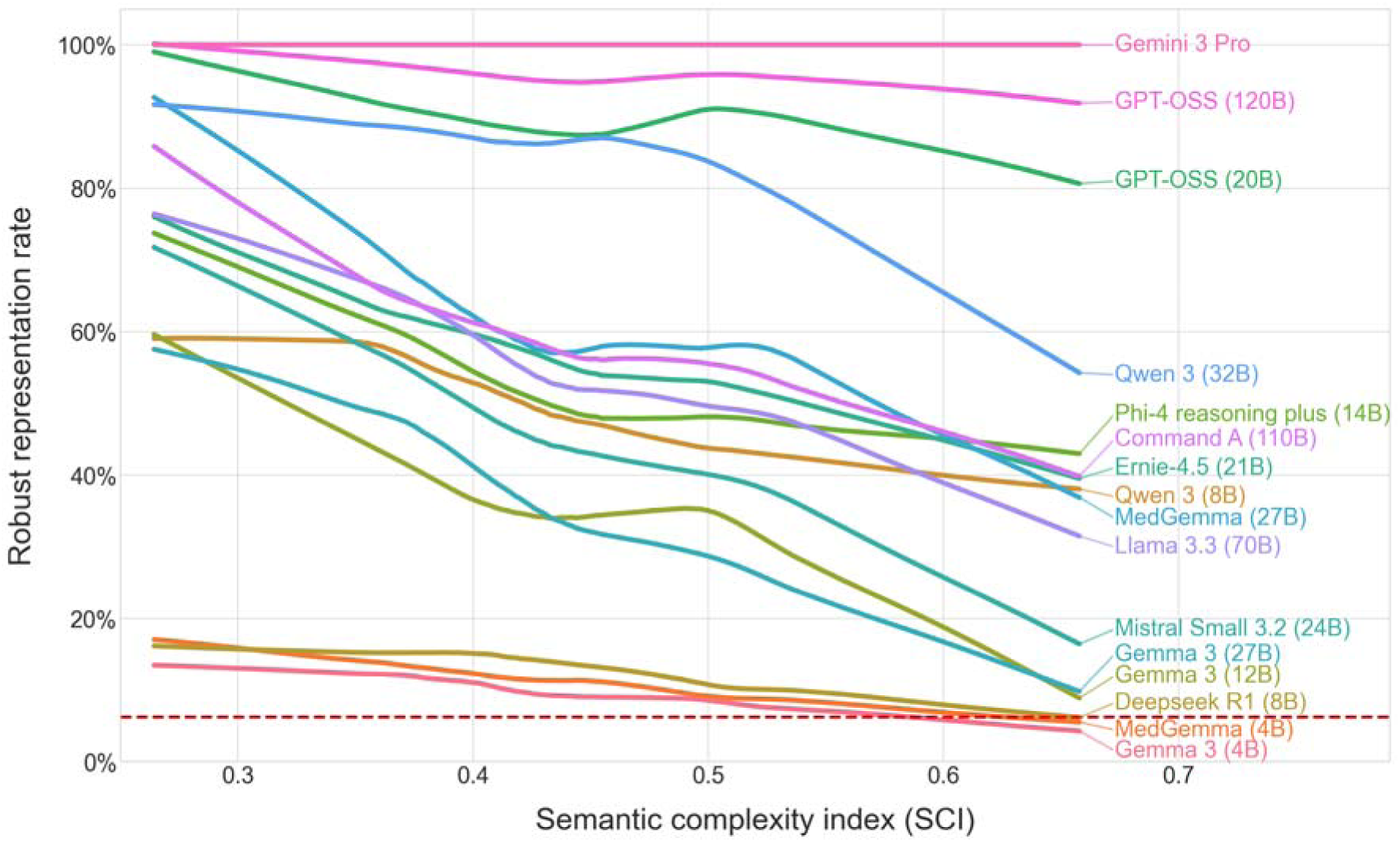
Model performance relative to semantic complexity of terms. LOWESS regression curves illustrate the conditional mean accuracy of each model against the Semantic Complexity Index (SCI). Curves were calculated using a smoothing fraction of 0.66 to estimate non-linear performance trajectories across the spectrum of term complexity. The red dashed line indicates the chance performance level (6.25%).

The highest-performing models, GPT-OSS (120B) and the reference frontier model Gemini 3 Pro, demonstrated ’complexity invariance’, maintaining pass rates exceeding 91.8% even as the complexity (SCI) increased from 0.26 to 0.66. In contrast, most models in the 12B to 110B range exhibited a strong negative relationship between complexity and representational robustness. While these models achieved robust performance on low-complexity terms, they suffered precipitous declines at the upper limits of the SCI. For example, Qwen 3 (32B) dropped from 91.7% to 53.8%, Command A (110B) fell from 81.8% to 39.8%, and Llama 3.3 (70B) declined from 70.6% to 30.8%.

A notable exception to this pattern was GPT-OSS (20B), which maintained relatively high stability (declining only from 98.2% to 80.7%), and outperforming models five times its size on high-complex terms.

### Medical fine-tuning enhances representational robustness in 27B but not 4B models independent of semantic complexity

To assess the impact of medical fine-tuning, we compared standard Gemma 3 models with their fine-tuned MedGemma variants for both 4B and 27B models. The 4B parameter models showed no significant benefit from fine-tuning (14.7% +/- 2.5% vs 15.7% +/- 2.6%; P=0.67 for). In contrast, the 27B scale demonstrated a significant benefit from fine-tuning, with the robust representation rate increasing from 38.2% (+/- 3.5%) in the base model to 62.7% (+/-3.5%) in the MedGemma variant (P < 0.0001 for difference).

We considered whether this performance uplift was concentrated in either low- or high-complexity terms, categorised by a median split of the SCI. While fine-tuning in the 27B model yielded numerically higher gains for high-complexity than low-complexity terms (+27.5% vs +21.3%), the lack of a significant interaction (*P*=0.22) indicates that medical fine-tuning provided a consistent performance benefit across the range of term complexity.

## DISCUSSION

This work presents three key findings regarding the robustness of medical terminology representations in locally deployable language models. First, neither parameter size nor specialist medical fine-tuning guarantees representational robustness. Notably, a mid-sized generalist model (GPT-OSS 20B) demonstrated greater robustness than both a larger, medically fine-tuned, model (27B) and far larger (70–110B) general-purpose architectures. Second, robustness of representations is conditional on terminological complexity. High performance on simple terms often masks precipitous failures when processing complex medical terminology. Third, representational robustness is subdomain-dependent, with models demonstrating significantly greater robustness for terminology linked to diagnoses (e.g. *Miller-Fisher syndrome*) than anatomy (e.g., *Broca’s area*) or clinical features (e.g., *Tongue fasciculation*).

Collectively, these findings challenge as heuristic that larger or medically fine-tuned LLMs are globally superior for clinical deployment. While robustness broadly adhered to neural scaling laws^35^, we observed significant exceptions where size did not equate to performance. For most local LLMs, robustness was highly conditional on terminological complexity; thus, high performance on general medical tests may not transfer to specialised settings ^36^.

To be clinically viable, an LLM must demonstrate ‘complexity invariance’, reliable performance regardless of a term’s societal prominence, ambiguity, or rarity. Outside of frontier models, only GPT-OSS 120B and 20B achieved this. Notably, GPT-OSS 20B maintained scores above 80% across all complexities, outperforming models five times its size by 40–50% on complex terms. This underscores that architectural optimization and training quality can be more influential than raw parameter count^37^.

The SCI helps match models to use cases by quantifying representational difficulty. Developers can use it to stratify clinical tasks by risk; lightweight models may suffice for low-complexity terminology, but complex tasks require the robustness found only in frontier models or the GPT-OSS series.

We also found that medical fine-tuning is constrained by architectural capacity. The 4B model hit a performance floor, whereas the 27B model utilised fine-tuning for consistent gains across all complexities. Consequently, local deployment strategies should prioritise fine-tuning only for models large enough to effectively integrate specialised knowledge.

This study had limitations. First, we did not assess multimodal LLM capabilities (e.g. combining text with medical imaging data). However, we argue that a model’s capacity to interpret multimodal data relies on a robust linguistic and logical foundation; if the underlying term representations are fragile, any downstream processing of those terms carry inherent risk regardless of input modality.

Second, we evaluated medical terms in a low-context environment rather than embedded within complex clinical narratives (e.g. a discharge summary where a diagnosis might be mentioned alongside negated symptoms and competing differential diagnoses). While this reduces ecological validity, it provided necessary experimental control; a model incapable of robustly representing a term in isolation cannot be trusted to interpret that term within the noise of unstructured documentation^36^.

Third, our focus here on term-level representational robustness differs from benchmarks that prioritise higher-level clinical reasoning^15–17^. Recent evidence suggests that high scores on medical leaderboards often mask ’shortcut learning’ and brittleness, where models pass tests using pattern-matching tricks rather than genuine medical understanding^38^. By evaluating robustness at this foundational level, we are testing the ’atomic’ stability of their representation of medical terms often presupposed by current benchmarks. We argue that this foundational robustness is an essential prerequisite for safety; without it, high-level reasoning is susceptible to catastrophic failure in the complex, real-world scenarios that standardised examinations may fail to detect.

## Conclusion

While local LLM deployment offers distinct privacy advantages^1^, these benefits must be weighed against demonstrated representational limitations. Our findings indicate that neither parameter scale nor domain-specific fine-tuning are reliable proxies for the robustness of medical term representations. Because the integrity of a clinical AI system is predicated on the stability of its core medical representations, a fragile foundation introduces a risk of unpredictable failure when the system is confronted with the atypical scenarios common in clinical practice. Consequently, we propose that verifying term-level robustness, using complexity-aware frameworks like the SCI, be considered a prerequisite for clinical adoption. Such rigorous validation ensures that as we move toward local deployment, these models remain reliable not only for routine tasks but also within the nuanced edge cases of modern medicine.

## Data and code availability

The dataset of 250 neurological term triplets used in this study and all python code, including the full text of LLM prompts will all be made freely available upon publication in a dedicated repository within https://github.com/stepdaug.

## Data Availability

The dataset of 250 term triplets used in this study and all python code, including the full text of LLM prompts will all be made freely available upon publication in a dedicated repository within https://github.com/stepdaug.

## Acknowledgements

SDA is supported by a UK National Institute for Health and Care Research (NIHR) Clinical Lectureship and acknowledges infrastructure support for this research from the NIHR Imperial Biomedical Research Centre (BRC).

NSNG acknowledges support from Alzheimer’s Society and Imperial NIHR BRC.

GS, NIHR Advanced Fellow, NIHR302971 is funded by the NIHR for this research project. The views expressed in this publication are those of the author and not necessarily those of the NIHR, NHS or the UK Department of Health and Social Care

The authors also acknowledge MRC equipment support via the UK Dementia Research Institute and are grateful to Payam Barnaghi for assistance in accessing this.

## Author Contributions

SDA conception, design, data analysis, manuscript first draft and review. NSN stimuli and manuscript review. PB infrastructure support and manuscript review. GS design, stimuli and manuscript review.

