## Supplementary materials for "On the robustness of medical term representations in locally deployable language models"

| **Model Name** | **Developer** | **Parameters** | **Type** | **Access** |
| --- | --- | --- | --- | --- |
| Gemini 3 Pro | Google | Undisclosed (Est >1T) | General | Proprietary |
| GPT-OSS (120B) | OpenAI | 117B (5.1B active) | General | Open Weights |
| Command A (110B) | Cohere | 111B | General | Open Weights |
| Llama 3.3 (70B) | Meta | 70B | General | Open Weights |
| Qwen 3 (32B) | Alibaba | 32.5B | General | Open Weights |
| MedGemma (27B) | Google | 27B | Medical | Open Weights |
| Gemma 3 (27B) | Google | 27B | General | Open Weights |
| Mistral Small 3.2 (24B) | Mistral AI | 24B | General | Open Weights |
| Ernie-4.5 (21B) | Baidu | 21B | General | Open Weights |
| GPT-OSS (20B) | OpenAI | 21B (3.6B active) | General | Open Weights |
| Phi-4 Reasoning+ (14B) | Microsoft | 14B | General | Open Weights |
| Gemma 3 (12B) | Google | 12B | General | Open Weights |
| DeepSeek R1 (8B) | DeepSeek | 8B | General | Open Weights |
| Qwen 3 (8B) | Alibaba | 8B | General | Open Weights |
| MedGemma (4B) | Google | 4B | Medical | Open Weights |
| Gemma 3 (4B) | Google | 4B | General | Open Weights |

**Table S1: Technical specifications and architectural characteristics of evaluated Large Language Models.**

Overview of the 15 models included in the comparative analysis and the reference frontier model (Gemini 3 Pro), listed in descending order of parameter scale. Developer indicates the entity responsible for pre-training. Parameters refers to the total parameter count; for Mixture-of-Experts (MoE) architectures (e.g. GPT-OSS), total parameters are listed, with active parameter details noted where applicable. Type distinguishes between general-purpose foundation models and those explicitly fine-tuned on biomedical corpora. Access classifies models as either proprietary (API-gated) or Open Weights (available for local deployment).

**Supplementary Appendix**

**LLM prompts**

To ensure the detected associations were not artifacts of specific prompt phrasing, we tested the LLMs across three diverse reasoning frameworks (clinical, structural, and semantic) to verify the stability of the medical representations.

Prompt 1:

System Prompt
You are a Senior Neurologist. Determine if the first finding guarantees the second.

Return 'true' only if the presence of the First Entity medically necessitates the Second Entity without exception.
Return 'false' if the First Entity is a broader concept that does not strictly imply the specific Second Entity (Reverse Implication).
Return 'false' for statistical associations or comorbidities.

Reply with exactly one word: 'true' or 'false'.

User Prompt
First Entity: "{term_if}"
Second Entity: "{term_then}"

Does the First guarantee the Second?

Prompt 2:

System Prompt
You are a Senior Medical Ontologist. Evaluate the hierarchical relationship between medical concepts.

Return 'true' if EITHER of the following conditions is met:

1. IS-A Relationship: Term A is a specific subtype or instance of Category B.
2. PART-OF Relationship: Term A is part of Category B.
3. INDICATOR-OF Relationship: Term A indicates the definite presence of Category B.

Return 'false' if:

- Term A is the broader category.
- Term A is merely associated with B (e.g., risk factors or loose correlations).
- Term A is distinct from B.

Output strictly: 'true' or 'false'.

User Prompt
Term A: "{term_if}"
Category B: "{term_then}"

Is A a subtype, part, or definite indicator of B?

Prompt 3:

System Prompt
You are a Medical Semantic Auditor. Determine if Term B is an intrinsic definitional requirement of Term A.

Apply this test: "By medical definition, is Term A inherently classified as a type, component or direct indicator of Term B?"

Output strictly: 'true' or 'false'.

User Prompt
Term A: "{term_if}"
Term B: "{term_then}"

Is B inherently required to define A?
